# Alcohol Use Disorder-Associated DNA Methylation in the Nucleus Accumbens and Dorsolateral Prefrontal Cortex

**DOI:** 10.1101/2024.01.17.23300238

**Authors:** Julie D. White, Melyssa S. Minto, Caryn Willis, Bryan C. Quach, Shizhong Han, Ran Tao, Amy Deep-Soboslay, Lea Zillich, Shaunna L. Clark, Edwin J. C. G. van den Oord, Thomas M. Hyde, R. Dayne Mayfield, Bradley T. Webb, Eric O. Johnson, Joel E. Kleinman, Laura J. Bierut, Dana B. Hancock

**Affiliations:** GenOmics and Translational Research Center, RTI International; Lieber Institute for Brain Development (LIBD); Department of Genetic Epidemiology in Psychiatry, Central Institute of Mental Health, Medical Faculty Mannheim, Heidelberg University, Mannheim, Germany; Department of Psychiatry & Behavioral Sciences, Texas A&M University; Center for Biomarker Research and Precision Medicine, Virginia Commonwealth University; Waggoner Center for Alcohol and Addiction Research, The University of Texas at Austin; Fellow Program, RTI International; Department of Psychiatry, Washington University School of Medicine

**Keywords:** epigenome-wide association study, substance use, postmortem human brain, gene regulation

## Abstract

**Background:** Alcohol use disorder (AUD) has a profound public health impact. However, understanding of the molecular mechanisms underlying the development and progression of AUD remain limited. Here, we interrogate AUD-associated DNA methylation (DNAm) changes within and across addiction-relevant brain regions: the nucleus accumbens (NAc) and dorsolateral prefrontal cortex (DLPFC).

**Methods:** Illumina HumanMethylation EPIC array data from 119 decedents of European ancestry (61 cases, 58 controls) were analyzed using robust linear regression, with adjustment for technical and biological variables. Associations were characterized using integrative analyses of public gene regulatory data and published genetic and epigenetic studies. We additionally tested for brain region-shared and -specific associations using mixed effects modeling and assessed implications of these results using public gene expression data.

**Results:** At a false discovery rate ≤ 0.05, we identified 53 CpGs significantly associated with AUD status for NAc and 31 CpGs for DLPFC. In a meta-analysis across the regions, we identified an additional 21 CpGs associated with AUD, for a total of 105 unique AUD-associated CpGs (120 genes). AUD-associated CpGs were enriched in histone marks that tag active promoters and our strongest signals were specific to a single brain region. Of the 120 genes, 23 overlapped with previous genetic associations for substance use behaviors; all others represent novel associations.

**Conclusions:** Our findings identify AUD-associated methylation signals, the majority of which are specific within NAc or DLPFC. Some signals may constitute predisposing genetic and epigenetic variation, though more work is needed to further disentangle the neurobiological gene regulatory differences associated with AUD.

## Introduction

Alcohol use disorder (AUD), characterized by tolerance, withdrawal and/or craving with cessation of use, and continued alcohol use despite adverse social or health consequences, is a highly prevalent neuropsychiatric disease that affected approximately 28.6 million adults in the United States in 2021^1^. Given the profound impact on individuals and society, research in AUD-relevant brain regions is critical to better understand the molecular mechanisms underlying the development and progression of AUD.

Epigenetic modifications, particularly DNA methylation (DNAm), are central to the interplay between genetic variation and environmental influences and may represent predisposing features to, or consequences of, alcohol use. This study presents an epigenome-wide association study (EWAS) of AUD in the nucleus accumbens (NAc) and dorsolateral prefrontal cortex (DLPFC) of postmortem brain comparing decedents with a history of AUD and well-matched controls. The NAc underlies cognitive processing of motivation, pleasure, and reward/reinforcement learning and is primarily implicated in the binge/intoxication stage of addiction, with a secondary role in the withdrawal/negative affect stage^2–4^. The DLPFC controls inhibition of impulsive responses, cognitive flexibility, planning, and abstract reasoning, and is involved in the preoccupation/anticipation stage of addiction^2^.

Prior EWAS of alcohol-related behaviors in human postmortem brain have total sample sizes ranging from 46 to 96. Most studies have used the Illumina 450K or EPIC (∼850K CpG sites) arrays, with AUD-associated differential DNAm found within Brodmann Area 9 (BA9; including parts of the dorsolateral and medial prefrontal cortex)^5,6^, the precuneus^7^, Brodmann Area 10 (anterior-most portion of the prefrontal cortex)^8^, caudate nucleus^9^, and ventral striatum^9^. Another recent study assayed both 5mC methylation and hydroxymethylation from the BA10 region of 50 decedents using methyl-CG binding domain sequencing, resulting in near-complete coverage of the 28 million possible methylation sites in the genome; no methylome-wide significant sites were found^10^. No overlapping sites have been reported across these studies, which is not surprising given the highly context-specific nature of DNAm, different analytic strategies, and small sample sizes.

The present study examines epigenome-wide AUD-associated DNAm in brain using the largest sample size to date, N = 119 decedents non-overlapping those included in the efforts summarized above. We assayed DNAm from the NAc and DLPFC of the same decedents to shed light on AUD-associated DNAm variation that is shared across and unique to these brain regions. Shared DNAm associations across brain regions may indicate AUD-associated genomic variation influencing DNAm or brain-wide responses to AUD. Alternatively, AUD-associated DNAm that is specific to a single brain region may indicate a more localized neurobiological AUD signal. We consider both possibilities for this study, which reveals intricate relationships between AUD and DNAm.

## Methods and materials

### Human postmortem NAc and DLPFC samples

Postmortem human NAc and DLPFC tissues were obtained at autopsy by the Lieber Institute for Brain Development Human Brain Repository^11–14^. Decedents with brain trauma, metastatic brain cancer, neuritic pathology, neurodegenerative diseases, HIV/AIDS, hepatitis, or other communicable diseases were excluded. At the time of tissue donation, the legal next-of-kin completed a 39-item questionnaire to obtain medical, social, psychiatric, substance use, and treatment histories. Retrospective clinical diagnostic reviews combined data from autopsy records, toxicology testing, forensic investigations, neuropathological examinations, questionnaires, psychiatric/substance abuse treatment record reviews, and/or supplemental family informant interviews (when possible and needed). All data were compiled in detailed psychiatric narratives which were independently reviewed by two board-certified psychiatrists to determine lifetime psychiatric diagnoses according to the Diagnostic and Statistical Manual of Mental Disorders, Fifth Edition (DSM-5), criteria.

We defined AUD cases as decedents with a lifetime history of two or more DSM-5 AUD symptoms within a 12-month period, with or without positive postmortem ethanol toxicology. AUD controls had no lifetime history of DSM-5 AUD symptoms and postmortem ethanol toxicology of less than 0.06 g/dL. We matched^15^ AUD cases and controls based on age, sex, smoking status (based on cotinine or nicotine biomarker and next-of-kin reporting)^11^, major depression diagnosis (MDD), and other drug indicators (mainly selective serotine reuptake inhibitors [SSRIs]). Decedents with MDD were defined as those with a lifetime history of five or more DSM-5 MDD symptoms persisting for two weeks or longer. Decedents meeting DSM-5 criteria for any other substance use or psychiatric disorder, besides AUD and MDD, were excluded. We included only decedents reported as white, aged 25 years old or older, and with no detectable opioids to minimize confounding effects. Sample characteristics are displayed in Table 1.

**Table 1.**
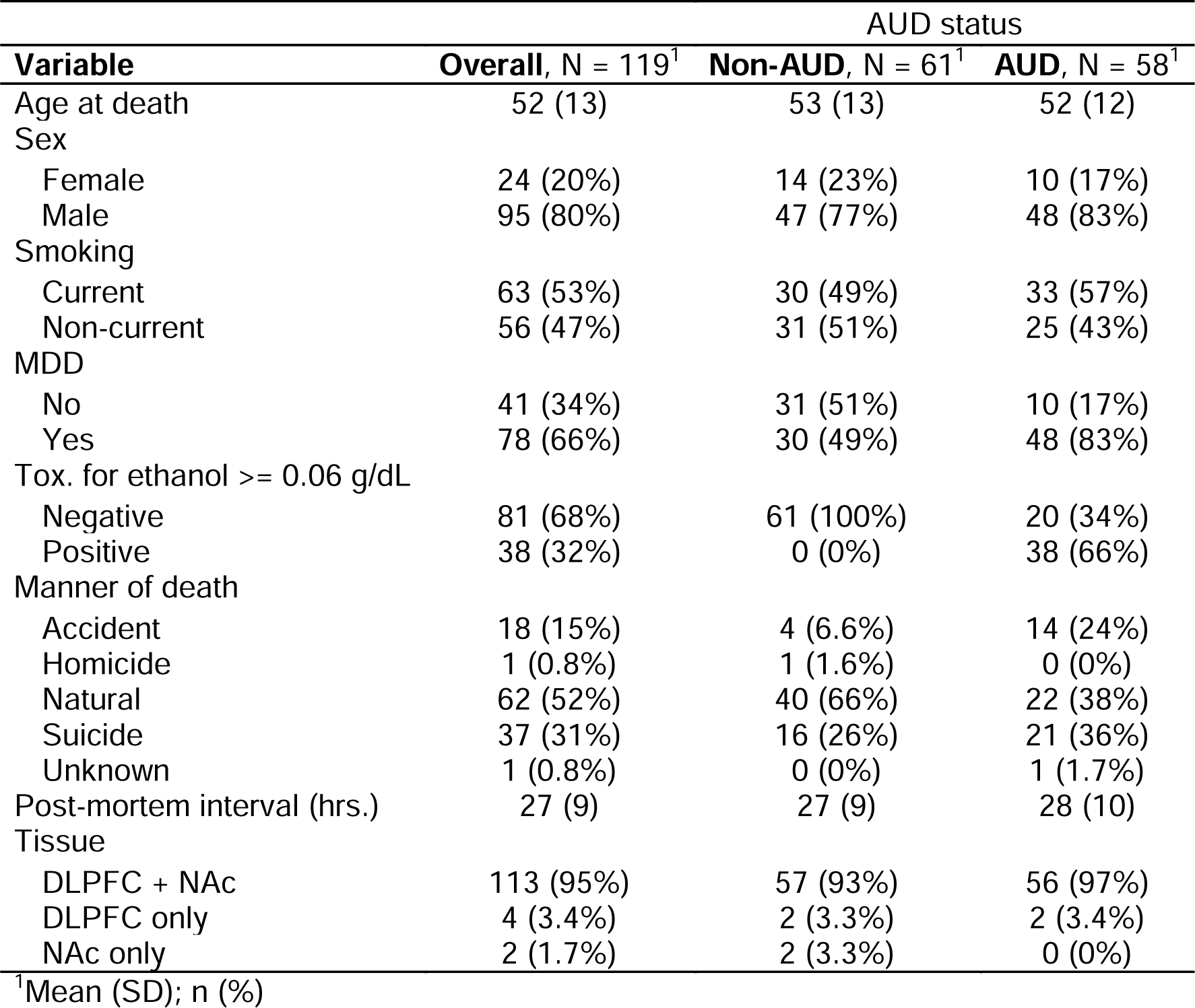
Description of samples used for analysis (N = 119).

| Variable | Overall, N = 119<sup>1</sup> | AUD status |  |
| --- | --- | --- | --- |
|  |  | Non-AUD, N = 61<sup>1</sup> | AUD, N = 58<sup>1</sup> |
| Age at death | 52 (13) | 53 (13) | 52 (12) |
| Sex |  |  |  |
| Female | 24 (20%) | 14 (23%) | 10 (17%) |
| Male | 95 (80%) | 47 (77%) | 48 (83%) |
| Smoking |  |  |  |
| Current | 63 (53%) | 30 (49%) | 33 (57%) |
| Non-current | 56 (47%) | 31 (51%) | 25 (43%) |
| MDD |  |  |  |
| No | 41 (34%) | 31 (51%) | 10 (17%) |
| Yes | 78 (66%) | 30 (49%) | 48 (83%) |
| Tox. for ethanol $\geq 0.06$ g/dL | | | |
| Negative | 81 (68%) | 61 (100%) | 20 (34%) |
| Positive | 38 (32%) | 0 (0%) | 38 (66%) |
| Manner of death |  |  |  |
| Accident | 18 (15%) | 4 (6.6%) | 14 (24%) |
| Homicide | 1 (0.8%) | 1 (1.6%) | 0 (0%) |
| Natural | 62 (52%) | 40 (66%) | 22 (38%) |
| Suicide | 37 (31%) | 16 (26%) | 21 (36%) |
| Unknown | 1 (0.8%) | 0 (0%) | 1 (1.7%) |
| Post-mortem interval (hrs.) | 27 (9) | 27 (9) | 28 (10) |
| Tissue |  |  |  |
| DLPFC + NAc | 113 (95%) | 57 (93%) | 56 (97%) |
| DLPFC only | 4 (3.4%) | 2 (3.3%) | 2 (3.4%) |
| NAc only | 2 (1.7%) | 2 (3.3%) | 0 (0%) |

<sup>1</sup>Mean (SD); n (%)

### DNAm data, quality control (QC), and pre-processing

DNA was extracted from NAc and DLPFC (BA 46/9), following Lieber’s protocol^16,17^. DNAm was measured using the Illumina HumanMethylation EPIC array. Data quality assessment and pre-processing were conducted using minfi (v1.40.0)^18^. We excluded probes with low call rate, existence of a common single nucleotide polymorphism in the extension site, cross-reactive probes^19,20^, probes mapped to sex chromosomes based on hg19 annotation, and probes with low bead count. Probes were quantile normalized. No samples met exclusion criteria for low call rate (detection *p* > 0.01 for >1% of probes), mismatched sex, and methylated vs. unmethylated values <10. One assay chip produced outliers based on DNAm-derived principal components (PCs), resulting in the removal of 5 samples for NAc and 3 samples for DLPFC. CpGs were later annotated to genomic locations based on GRCh38 / hg38 information from Zhou, et al.^21^, (https://zwdzwd.github.io/InfiniumAnnotation, accessed August 2022). After all QC and processing steps, we analyzed 769,135 CpG sites from 115 NAc samples and 767,700 CpG sites from 117 DLPFC samples (Table 1; Table S1).

### AUD EWAS analyses

We used robust linear regression to compare DNAm between AUD cases and controls, with methylation M-values as the outcome. We calculated robust standard errors using White’s estimator in the sandwich package (v3.0.2)^22,23^. Analyses were adjusted for MDD (case vs. non-case), age at death, sex, smoking (current vs. not), five DNAm-derived PCs, plate (P1 vs. P2, P1 vs. P3), and row position on methylation chip (rows 1-4 vs. 5-8). Cell type composition references for human brain DNAm data are unavailable for NAc. Therefore, for both brain regions, PCs calculated from the DNAm data were included to correct for technical artifacts and cellular heterogeneity. Results were corrected for inflation using the empirical null distribution estimation method implemented in bacon (v1.18.0)^24^, and lambda values were calculated on bacon-corrected p-values. Meta-analysis of bacon-adjusted p-values for the 766,095 CpGs intersecting both analyses was performed using METAL, with correction for sample overlap^25^. Similarity of effects across brain regions was assessed using I^2^ heterogeneity statistics. Significance was assessed at false-discovery rate (FDR) ≤ 0.05^26^.

### AUD-associated CpG tests against postmortem ethanol toxicology

Of the 58 decedents with AUD, 38 (66%) had positive postmortem ethanol toxicology. To assess if the significant AUD-associated CpGs displayed differential methylation by recent alcohol exposure, we used robust linear regression of methylation M-values on ethanol toxicology status (positive [≥0.06 g/dL] vs. negative [<0.06 g/dL]). This analysis was performed within AUD cases for the significant CpGs from the NAc, DLPFC, or meta-analysis and corrected for postmortem interval (hours) and all other covariates used in the AUD case/control EWASs. Significance was assessed at FDR ≤ 0.05^26^.

### Enrichment tests

Gene set enrichment for known pathways was tested using functions from missMethyl (v1.32.0)^27^. From each EWAS (NAc, DLPFC, meta-analysis), we supplied gsameth with a list of significant CpGs, all tested CpGs for that analysis, and gene sets to test for enrichment. CpG-to-gene mapping was based on Zhou et al.^21^ after unique gene symbols were converted to Entrez gene ids using the AnnotationDbi (v1.60.0)^28^ and org.Hs.eg.db (v3.16.0)^29^ packages. Only gene symbols with an Entrez gene id were considered. We allowed a single CpG to be annotated to multiple genes.

Two-sided Fisher’s exact test for count data was used to test for enrichment of AUD-associated probes in CpG (islands, shelves, and shores) and genic (promoters, 5’UTRs, exons, introns, and 3’UTRs) contexts using the stats package (v4.2.2). The annotatr (v1.24.0)^30^ and LOLA (v1.28.0)^31^ packages were used to intersect CpG’s hg38 positions with CpG and genic locations, respectively, with a minimum overlap of 2. We additionally used LOLA to test for enrichment of significant CpG positions with ChIP-seq histone modification sites (H3K27ac, H3K27me3, H3K36me3, H3K4me1, H3K4me3, H3K9ac, and H3K9me3) derived from Roadmap Epigenomics Consortium epigenomes cultured from brain cells or tissues^32^. All Roadmap epigenomes tested were generated from non-diseased decedents of varying age and sex (Table S2).

### Concordance with published EWAS of AUD and alcohol consumption

We compared our results with previously published EWAS of AUD or alcohol consumption in two ways to capture probe-level and overall concordance between our results and those of previous publications. We selected previous EWAS with large sample sizes (defined below), results reported by probe or coordinates, and DNAm assayed from blood or brain tissues. Three selected studies tested for associations with alcohol abuse, dependence, or use disorder in brain tissues^7,9,10^ (n ≥ 46), and two tested for associations with alcohol consumption in blood^33,34^ (n > 5,000). First, we performed a “look-up” of previously reported sites in our results. For CpGs that were tested in one of our EWAS analyses and identified as significant in a previous study, or among the top 20 CpGs if no significant results were reported, we calculated new Benjamini-Hochberg FDR-corrected p-values and declared “look-up” statistical significance based on FDR ≤ 0.05.

Second, we requested the full summary statistics from Zillich *et al.*’s EWAS of AUD in five brain regions^9^ and Clark *et al.*’s BA10 whole genome methylation and hydroxymethylation AUD-association study^10^ to test for p-value enrichment in our study. These studies were selected because they had the most overlapping methylation sites with our study and tested DNAm in brain. For each combination of brain regions in Zillich *et al.* or methylation assays in Clark *et al.*, we intersected results and subsequently tested for enrichment of the lowest 1% of p-values from our study in the lowest 1% of p-values from the previous publication. This threshold balanced between including too few sites, which could reduce power, and including too many sites, which may lead to false positive results. Testing was performed using the enrichmentAnalysis function in the shiftR package (v1.5), with 10,000 permutations^35^.

### Concordance with genome-wide association study (GWAS) results for alcohol behaviors

We used stratified linkage disequilibrium score regression (LDSC; v1.0.1)^36,37^ to calculate partitioned heritability estimates and test for enrichment of alcohol-associated genetic loci in annotation windows of 5 kb, 10 kb, 100 kb, 250 kb, and 500 kb around significant CpGs from our NAc, DLPFC, and meta-analysis results. We relied on summary statistics from GWAS of drinks per week^38^ and DSM-IV alcohol dependence^39^, both performed in European-ancestry individuals. 1000 Genomes Phase 3 “EUR” reference data^40^ was used to calculate linkage disequilibrium. We additionally used gwasrapidd (v0.99.14)^41^ to search for addiction related associations previously reported in the GWAS catalog^42^ for our significant genes.

### Linear mixed-effects modeling of brain region-shared and -specific AUD-associated DNAm

We conducted a meta-analysis to identify CpGs with directionally consistent associations with AUD, albeit potentially with smaller effect sizes, across the two brain regions. CpGs that were significant in our NAc or DLPFC EWAS, but not in the meta-analysis, may have resulted from different methylation levels for that CpG in the two brain regions, or from associations that differed between AUD and methylation across the brain regions. To explicitly test these scenarios, we used linear mixed-effects modeling to test for differential CpG methylation by brain region, AUD, and a brain region × AUD interaction. CpGs associated with the AUD term have similar associations across NAc and DLPFC; CpGs associated with the brain region term have different methylation levels across the two; and CpGs associated with the AUD × brain region interaction have different associations with AUD depending on brain region (i.e., brain region-specific effects).

To perform this modeling, we used a two-step approach to model within-subject differences in DNAm across brain regions and between-subject differences in AUD status. First, DNAm from each brain region was separately regressed on within-subject technical variables (PCs, plate, and row position on methylation chip) and the resulting residuals were carried forward. Next, a linear mixed-effects model was implemented with the lme4 package (v1.1.33)^43^, with residualized M-values as the outcome, MDD, age at death, sex, and smoking as adjustment variables, and AUD, brain region, and AUD × brain region as our effects of interest. This model included a random effect of ID to account for sampling two brain regions from the same decedent.

Using these results, we also identified differentially methylated regions (DMRs) for AUD, brain region, and their interaction using the comb-p function in ENmix (v1.34.02)^44,45^. For DMRs comprised of at least 2 CpGs, significance was assessed at a Sidak p-value ≤ 0.05. DMRs were annotated to genes with the nearest TSS using the annotatePeak function in ChIPSeeker (v1.36.0)^46^, with the TxDb.Hsapiens.UCSC.hg38.knownGene (v3.2.2)^47^ and org.Hs.eg.db (v3.16.0)^29^ packages.

### Profiling of expression across brain regions using the Allen Human Brain Atlas

To assess if genes nearby brain region-shared and -specific AUD-associated DMRs also had similarly shared and specific gene expression patterns in non-AUD brains, we utilized microarray gene expression data from six postmortem brains provided by the Allen Human Brain Atlas (AHBA)^48^. We selected expression data from the middle frontal gyrus (MFG-i) region, which contains the DLPFC, and the NAc (labeled Acb in AHBA). For each microarray probe targeting a gene, or its synonym, annotated to a significant DMR from our linear mixed-effects modeling results, we performed a paired t-test for differences in expression across the two brain regions. The ABHA microarray platform was designed such that multiple probes for a gene mapped to different exons as much as possible^48^. Thus, we treated each probe as unique and did not collapse across gene expression probes. Technical replicates were not collapsed due to the wide range of replicate numbers per donor, per brain region (Table S3). Instead, we employed a bootstrap sampling approach with 1,000 iterations, where in each iteration one replicate per brain region from each donor was selected, and a paired t-test was performed. We rejected the null hypothesis of no difference in expression across brain regions if the median bootstrapped p-value was ≤ 0.05.

## Results

### EWAS of AUD

Figure 1 provides a flowchart of our data, analyses, and results. We identified 53 and 31 CpGs significantly associated with AUD in NAc (λ = 1.03) and DLPFC (λ = 1.02), which were annotated to 65 and 36 genes, respectively. When combining results across brain regions, our meta-analysis identified 31 CpGs associated with AUD (λ = 1.03). Ten meta-analysis significant CpGs overlapped with significant CpGs from the NAc or DLPFC EWAS, though none were significant in all three analyses (Figure 2). In total, the EWAS analyses resulted in 105 CpGs associated with AUD annotated to 120 unique genes, which we carried forward as our primary results (Table S4).

**Figure 1.**
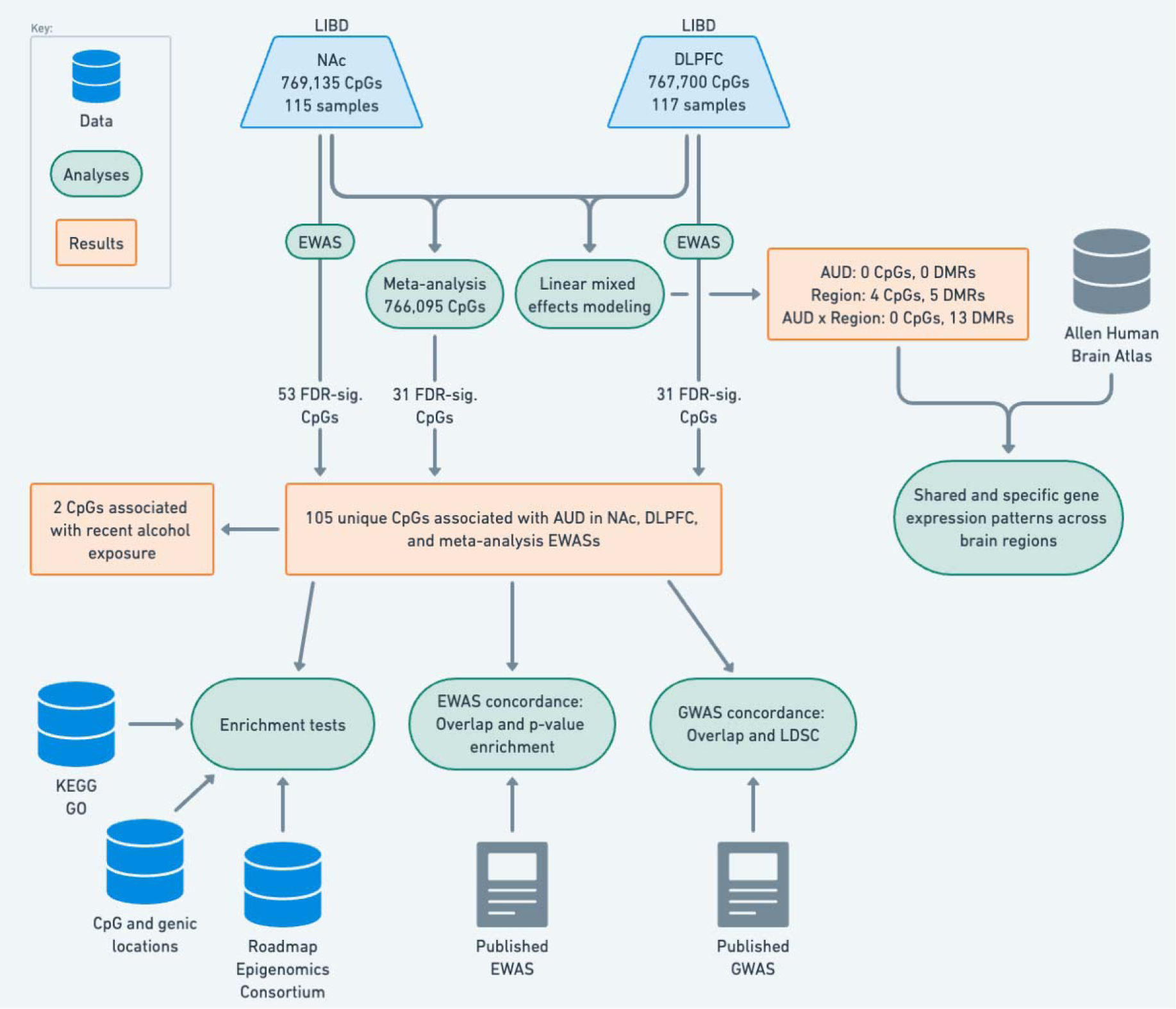
Overview of all data, analyses, and results.

**Figure 2.**
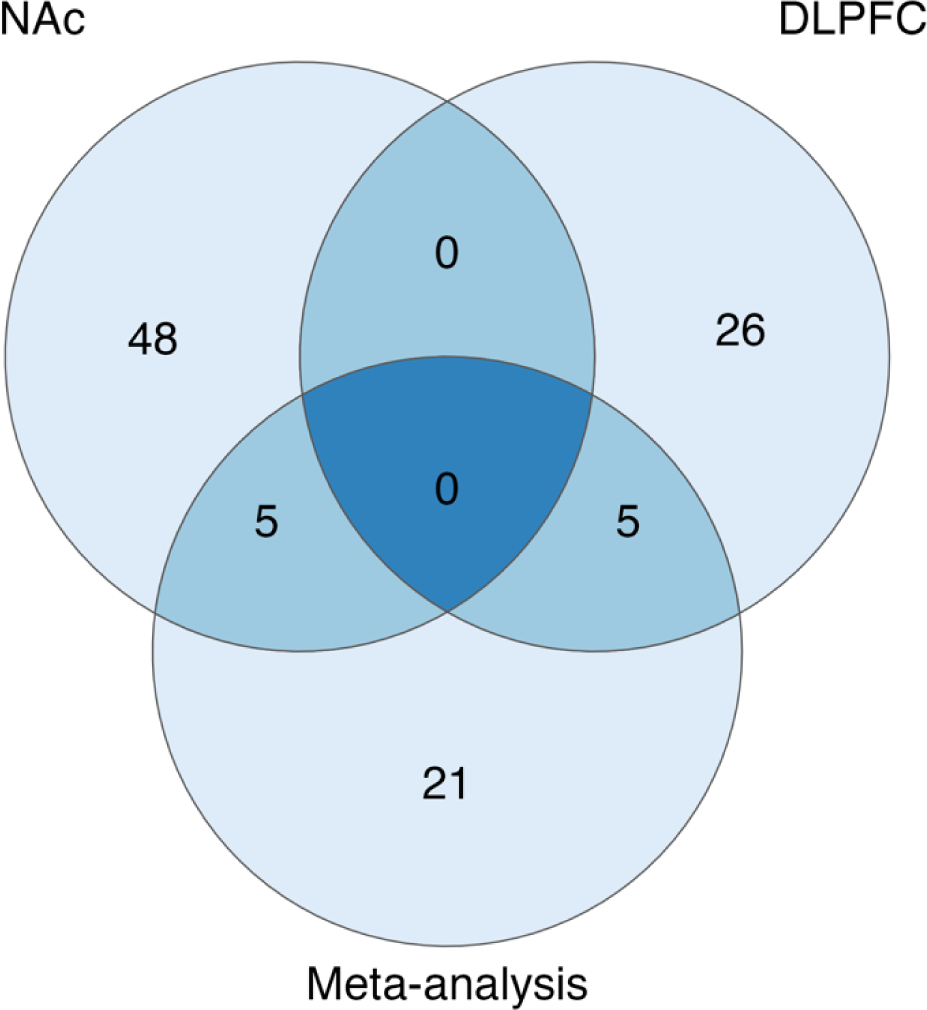
Overlap of significant probes from NAc and DLPFC EWAS analyses and the corresponding meta-analysis across brain regions.

### Differential methylation by recent alcohol exposure

A sensitivity analysis was conducted using decedents with AUD to test if recent alcohol exposure (defined by positive postmortem ethanol toxicology) influenced methylation among the 105 AUD-associated CpGs (Table S5). One CpG was significantly (FDR ≤ 0.05) associated with ethanol in the NAc (cg15747423; *UST*) and DLPFC analyses (cg25985151; *DNAI1*), respectively.

These results suggest that the CpG-AUD associations are largely robust to acute alcohol exposure around time of death.

### Enrichment tests

To characterize the AUD-associated CpGs, we performed several enrichment tests. We tested KEGG and GO pathway databases for enrichment using the 120 genes annotated for our AUD-associated CpGs. No pathways had FDR ≤ 0.05 (Table S6). In relation to CpG and genic locations, NAc AUD-associated CpGs were depleted in intergenic regions and enriched in islands, promoter regions, and 5’ UTRs (Figure 3; Table S7). No CpG or genic location enrichment/depletion was identified for the DLPFC AUD-associated CpGs. Meta-analysis AUD-associated CpGs were enriched in promoters, 5’UTRs, and exons (Figure 3B, Table S7). In the brain-derived cell and tissue Roadmap Epigenomics consortium epigenomes, significant CpGs from the NAc EWAS and meta-analysis were significantly enriched in H3K27ac, H3K9ac, and H3K4me3 marks (Figure S1, Table S7).

**Figure 3.**
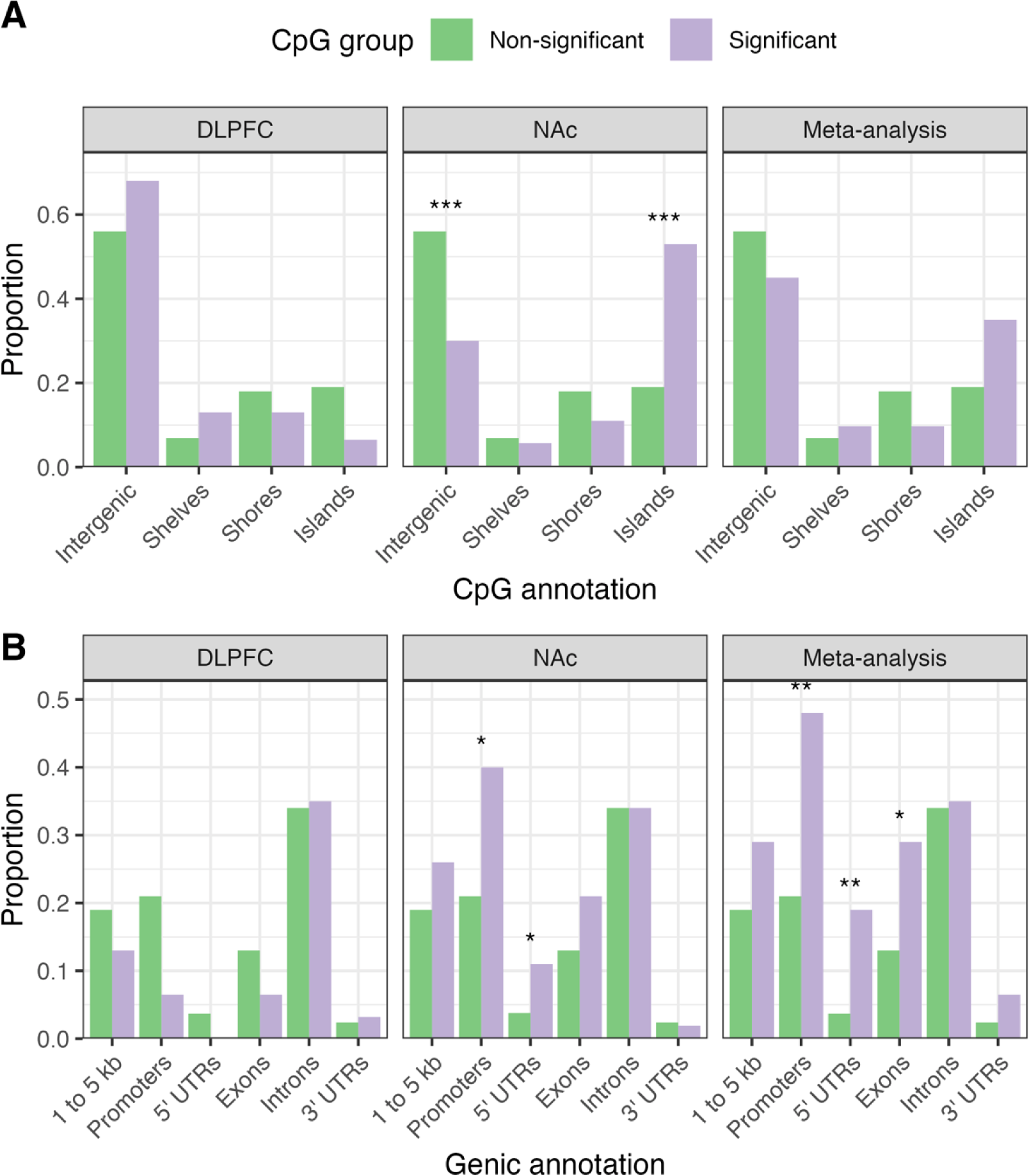
Enrichment of genomic features. Proportions of non-significant vs. significant CpGs were compared based on (A) CpG contexts and (B) gene-centric contexts. Stars represent significance of a two-sided Fisher’s exact test for count data, based on FDR-corrected p-values; p < 0.001 is ***, p < 0.01 is **, and p < 0.05 is *.

### Concordance of results with published EWAS of AUD and alcohol consumption

We compared our results to five published EWAS of abuse, dependence, or use disorder^7,9,10^ or alcohol consumption^33,34^ performed in either blood^33,34^ or brain^7,9,10^ (Table S8). Dugué *et al.* tested for associations with alcohol consumption in blood (N = 5,606). Of 1,237 CpGs available in our study out of their 1,415 significant CpGs^33^, three reached look-up significance in our NAc EWAS: cg03474926 (*RALGDS*), cg24678869 (*DENND4B*), and cg04162032 (*LYPD8*). Additionally, ten genes were annotated to probes that reached significance in our primary results and Dugué *et al.*, albeit for different probes (Figure 4). Lohoff *et al.* was the other large study that tested for alcohol consumption associations in blood (N = 8,161)^34^. While we did not identify any significant look-up results out of their 2,463 associated CpGs available in our study, 14 genes were annotated to significant probes in both our study and Lohoff *et al*., seven of which also overlapped with genes identified in Dugué *et al.*: *YARS1*, *RABGGTB*, *TRA2B*, *RREB1*, *RALGDS*, *CDH23*, and *ANKRD11* (Figure 4).

**Figure 4.**
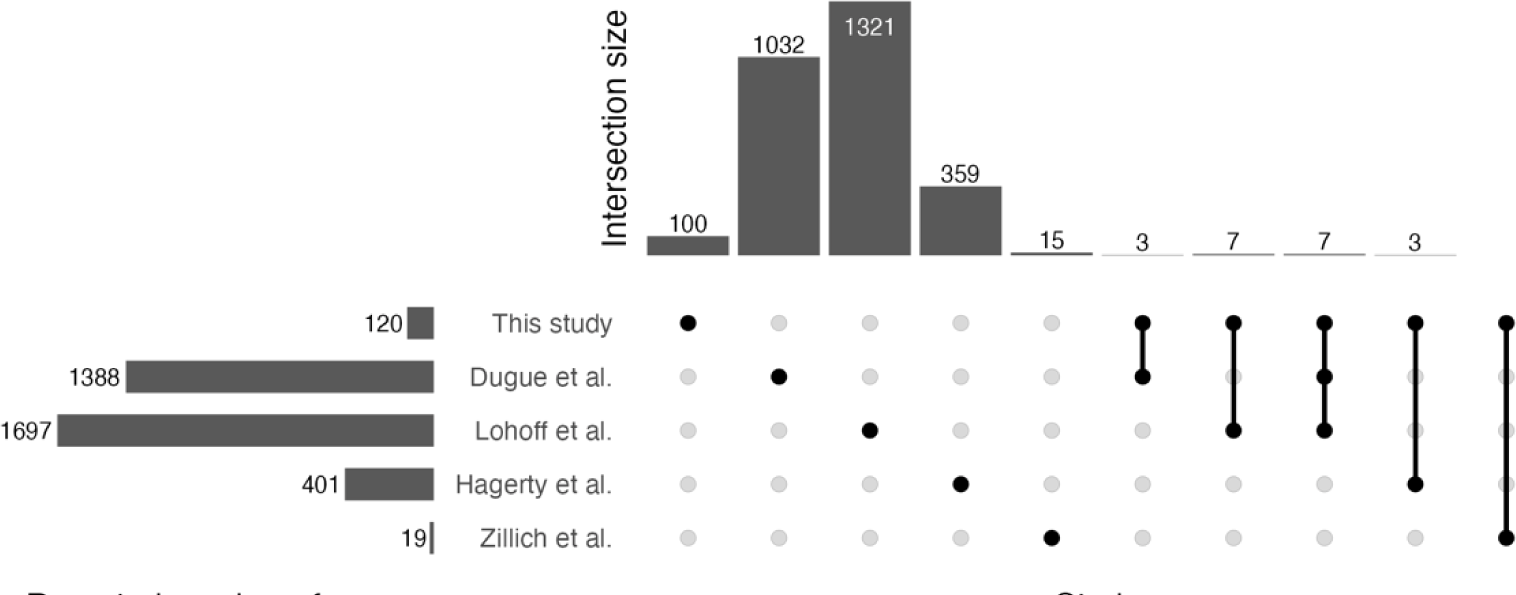
Gene-level concordance with previously published EWAS of AUD and alcohol consumption. This UpSet plot shows intersections between genes annotated to significant CpGs from our study (NAc, DLPFC, or meta-analysis) and previously published EWAS, considering only CpGs which passed the original publication’s significance threshold. Clark *et al.*’s results were not included, as no brain methylation or hydroxymethylation sites passed that study’s significance threshold. The comparison between our study and Zillich *et al.* did not have any overlapping genes.

Of the three studies that assayed methylation in brain tissues^7,9,10^, we identified one CpG with look-up significance and three annotated genes that overlapped with our results. cg00402668 (intergenic) reached look-up significance in the DLPFC EWAS and was among the top 20 sites with AUD-associated hydroxymethylation in BA10 from Clark *et al.*^10^. No CpGs reached look-up significance when comparing our results with Hagerty *et al.* (precuneus) and Zillich *et al.* (anterior cingulate cortex, DLPFC [BA9], putamen, caudate nucleus, and ventral striatum)^7,9^. On a gene level, three genes annotated to significant probes in our study overlapped with genes annotated to significant probes from Hagerty *et al.* (Figure 4; *CAPS2*, *PTPRN2*, and *SLIT3*)^7^. No genes overlapped in comparison to Zillich *et al*^9^.

Lastly, we compared the full summary statistics from our study with Clark *et al.*^10^ and Zillich *et al.*^9^, spanning six brain regions. When testing for enrichment of the top 1% of our results in the top 1% of Zillich *et al*.’s results, we identified significant enrichment for our NAc results in Zillich’s putamen and ventral striatum results but not for our DLPFC or meta-analysis results (Table S9)^9^. We did not observe significant enrichment between our results and those from Clark *et al.* (Table S9)^10^.

### Concordance with GWAS results

Stratified LDSC results did not indicate significant heritability enrichment of alcohol-associated genetic loci in varying genomic windows around significant CpGs from our primary analyses (Table S10). Twenty-three genes annotated to significant CpGs from our primary analyses were previously associated with GWAS results for substance use phenotypes, some of which were also reported in prior EWAS of the same phenotypes (Table 2).

**Table 2.**
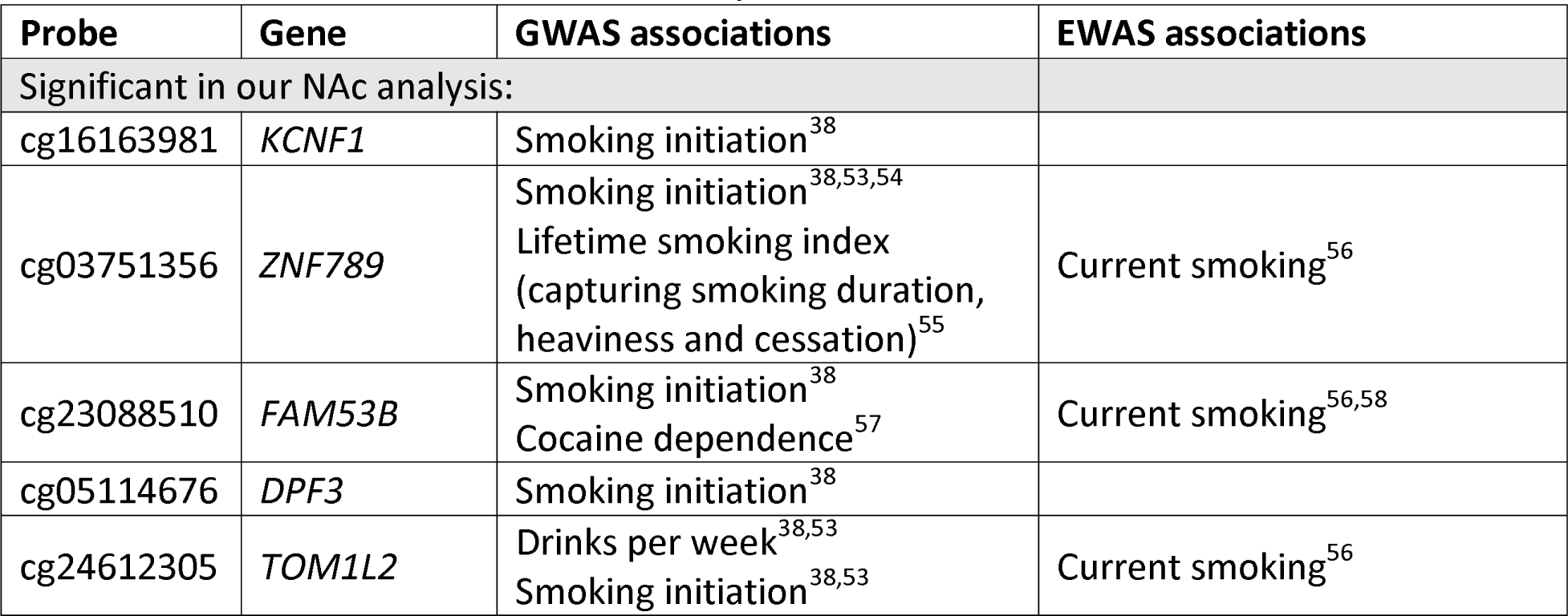
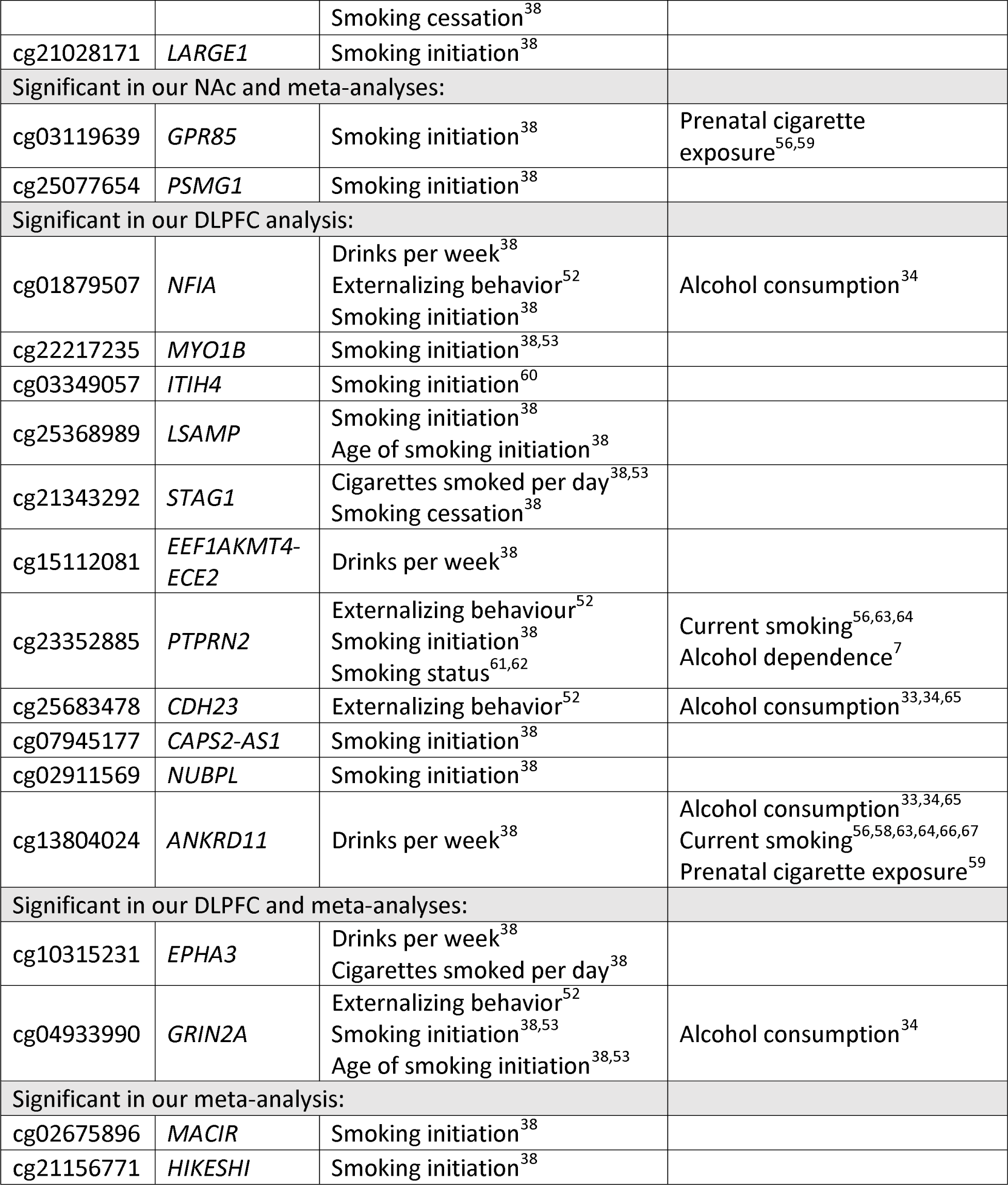
CpGs and annotated genes that were significant in our analyses and associated with alcohol or other substance use behaviors in published GWAS and EWAS.

### Shared vs. brain region-specific effects

CpGs that reached significance in the NAc and DLFPC analyses had very little similarity in effect sizes (Pearson’s r = 0.22; Figure S2A) and high I^2^ heterogeneity values, compared with CpGs that reached significance in the meta-analysis, which had much higher similarity of effect sizes (Pearson’s r = 0.93; Figure S2B) and much lower I^2^ heterogeneity values (Table S11, Figure S2C). Thus, the meta-analysis identified CpGs with similar directions and effect, while significant CpGs from the within brain region EWAS analyses were more dissimilar, as expected.

To explicitly test for brain region-specific or -shared associations with AUD, we used linear mixed-effects modeling to test for differential CpG methylation by brain region, AUD, and a brain region × AUD interaction. Four CpGs had FDR ≤ 0.05 for the region term, indicating different methylation profiles across brain regions (Table S12). No CpGs reached genome-wide significance for the AUD or AUD × region terms.

When testing DMRs, we identified five significantly associated with the brain region term and 13 significantly associated with the interaction term (Table S13). To test if genes annotated to these DMRs also had brain region-specific or -shared expression profiles, we used microarray expression from the AHBA MFG-i and Acb brain regions, representing the DLPFC and NAc, respectively. In total, 17 unique genes were annotated to significant DMRs, with 15 of these, or their synonyms, present in the AHBA data (4 from brain region term, 11 from interaction term). Three of the four brain region-annotated genes (75%) and two of the 11 interaction-annotated genes (18%) had expression probes with median paired t-test values ≤ 0.05, indicating different expression profiles in MFG-i and Acb brain regions (Figure S3; Table S13).

## Discussion

We report the largest EWAS of AUD in postmortem human brain to date. In within-brain region EWAS analyses, we identified 53 CpGs (65 genes) associated with AUD for the NAc and 31 CpGs (36 genes) associated with AUD in the DLPFC. No CpGs overlapped between the NAc and DLPFC analyses. When we meta-analyzed results across the two brain regions, accounting for sample overlap, we identified 31 CpGs significantly associated with AUD, ten of which overlapped with CpGs identified in the DLPFC or NAc EWAS. Only two of the 105 unique CpGs from these analyses had evidence for association with ethanol toxicology, indicating that our results were robust to recency of drinking.

We identified enriched overlap between significant CpGs from our NAc EWAS and meta-analysis results and H3K27ac, H3K9ac, and H3K4me3 histone marks in non-diseased brain cells and tissues. H3K27ac is a classic marker of active enhancers and promoters, and H3K4me3 marks are commonly associated with transcription activation in nearby genes. Given that we also identified enrichment in H3K9ac marks, typically associated with active promoters, but not in H3K4me1 marks, typically associated with gene enhancers, our results suggest promoter-specific regulation of nearby genes. This hypothesis is corroborated by our findings of significant enrichment for NAc and meta-analysis significant CpGs in islands, gene promoter regions, 5’UTR, and exon regions.

Prior EWAS of alcohol phenotypes in postmortem brain have not identified any genome-wide significant sites that overlapped across studies. Our results followed this trend. However, when we further compared our results to previous EWAS findings of alcohol-use phenotypes in blood and brain, we identified four CpGs that were among the top hits of a previous study and reached look-up FDR significance in one of our primary results. Concordance across studies was also higher when considered at the gene level. For example, seven genes (*YARS1*, *RABGGTB*, *TRA2B*, *RREB1*, *RALGDS*, *CDH23*, and *ANKRD11*) were implicated in our study and two prior blood-based EWAS of alcohol consumption, and three genes (*CAPS2*, *PTPRN2*, and *SLIT3*) were annotated to significant CpGs in a previous EWAS in brain and also observed in our study. Some concordance was also evident in the enrichment of top CpGs from our NAc results in results from EWAS of alcohol dependence in putamen and ventral striatum. The ventral striatum contains the NAc, and is proximal to the putamen, suggesting greater concordance for nearby brain regions with similar functions. However, we did not identify enrichment between our DLPFC results and previous EWAS in BA9^9^, which correspond anatomically, or BA10^10^, which is nearby. As with the prior comparisons, this lack of overlap could be due to different sampling, analytic strategies, and current sample sizes and statistical power.

Differing methylation levels across brain regions, regardless of disease status^49–51^, could also explain different associations with AUD across different brain regions. We formally tested this hypothesis to tease apart brain region-shared vs. -specific associations with AUD. Larger sample sizes would have benefited this analysis, as only four CpGs (all for the brain region term, suggesting different methylation levels across the regions) reached FDR significance. Nonetheless, we identified five DMRs for the brain region term and 13 DMRs with brain region-specific associations with AUD. We followed up this analysis by testing for expression differences between the MFG-i, which contains the DLPFC, and the Acb (NAc) in the Allen Human Brain Atlas. As expected, only two of the eleven genes annotated to DMRs significant for the interaction term (*i.e.*, brain region-specific effects) had evidence of different expression profiles across MFG-i and Acb, while three out of four genes annotated to the brain region-associated DMRs had evidence of different expression patterns across brain regions. These analyses suggest that while harmonized analytic strategies and increased statistical power may increase discovery and overlap of results among studies, human brain-derived, alcohol-related epigenetic associations should be considered in as specific locales as possible, as associations found in one brain region may not translate to another.

We did not identify enrichment for GWAS variants of alcohol behaviors in the genomic regions surrounding our significant CpGs, suggesting that these DNAm sites were largely not genetically driven factors that predispose individuals to alcohol traits. At the gene level, 23 genes were previously associated with addiction-related traits, many of which also had previous addiction-related EWAS associations. For example, *NFIA,* which encodes a member of the nuclear factor 1 (NF1) family of transcription factors. This gene was identified in our DLPFC analysis, a previous EWAS of alcohol consumption^34^, and in GWAS of drinks per week^38^, externalizing behaviors (including substance use)^52^, and smoking initiation^38^. Another gene, *GRIN2A*, encodes a member of the glutamate-gated ion channel protein family, and was identified in our DLPFC analysis and a previous alcohol consumption EWAS^34^. Variants in this gene were previously associated with externalizing behaviors, including substance use^52^, and with smoking initiation^38,53^. These converging associations could indicate that GWAS variants around these genes could impact methylation in the NAc and DLPFC specifically and potentially alter the key functions of these brain regions, contributing to the initiation of alcohol use.

Though this EWAS is the largest to date for AUD in postmortem human brain, statistical power remains limited, especially for complex models testing interactions and employing multiple testing correction. Additionally, this study’s focus on white decedents means that results may not be generalizable. We may also have reduced power due to the inclusion of decedents with a lifetime history of AUD, as opposed to an active AUD diagnosis at death, as some differentially methylated CpGs could have reverted to control levels if drinking had stopped for an extended period. Because we analyzed DNAm from bulk tissue, cell-type specific patterns for AUD-associated methylation were not captured. Single-cell sequencing would enable us to determine whether our brain region-shared and -specific DNAm changes are due to different cellular composition across these brain regions or if cells within each brain region also have distinct DNAm associations with AUD.

Despite these limitations, our unique study design allowed us to integrate and compare associations with AUD across two brain regions important to the addiction cycle. Altogether, our results suggest that the strongest signals associated with AUD are brain region-specific, helping to illuminate potential gene regulatory mechanisms within these brain regions that may regulate response to AUD. We also identified many associations annotated to genes previously implicated in GWAS of substance use, particularly for cigarette smoking. These converging associations could indicate genetic variants at these genes altering methylation and predisposing an individual to a generalizable addiction liability for alcohol and other substance use. There are several explanations for the associations we identified that did not overlap with prior GWAS, including differential methylation at these sites reflecting consequences of excessive alcohol intake that may help explain neuroplastic changes in response to AUD, not predisposing factors. Larger sample sizes, meta-analyses, and other integrative efforts will help clarify these relationships, promote further understanding of the molecular mechanisms underlying AUD, and identify therapeutic options to help individuals with AUD.

## Acknowledgements

The authors would like to gratefully acknowledge the generosity of the families of the decedents, who donated the brain tissue used in this study. We are also thankful for the many colleagues whose efforts have led to the donation and curation of postmortem tissue that makes this study possible.

## Disclosures

### Data availability

Illumina EPIC methylation data generated from this study has been deposited on GEO (GSE252501) and summary statistics for all association analyses are provided as supplemental tables on Figshare at doi: 10.6084/m9.figshare.24871662.

