## Supplemental Information for "Alcohol Use Disorder-Associated DNA Methylation in the Nucleus Accumbens and Dorsolateral Prefrontal Cortex"

**Table S1.** Quality control filters applied to CpG probes in DLPFC and NAc

| Criteria | DLPFC | | | NAc | | |
|---|---|---|---|---|---|---|
| | **Before** | **After** | **N CpGs removed** | **Before** | **After** | **N CpGs removed** |
| Probes with low call rate (detection p > 0.01 in >5% of samples) | 866,238 | 864,497 | 1,741 | 866,238 | 864,342 | 1,896 |
| Probes with a single nucleotide polymorphism with minor allele frequency > 0.01 in the extension site | 864,497 | 834,677 | 29,820 | 864,342 | 834,524 | 29,818 |
| Cross-reactive probes[1,2] | 834,677 | 793,776 | 40,901 | 834,524 | 793,627 | 40,897 |
| Probes mapped to sex chromosomes based on hg19 | 793,776 | 776,159 | 17,617 | 793,627 | 776,008 | 17,619 |
| Bead count <3 in >5% of samples | 776,159 | 770,232 | 5,927 | 776,088 | 771,667 | 4,421 |
| 'ch' probes | 770,232 | 767,719 | 2,513 | 771,667 | 769,154 | 2,513 |
| **Final probe count at QC stage** | | **767,719** | | | **769,154** | |
| Probes mapped to sex chromosomes based on hg38 | 767,719 | 767,700 | 19 | 769,154 | 769,135 | 19 |
| **Final probe count for results interpretation** | | **767,700** | | | **769,135** | |

**Table S2.** Information on Roadmap Epigenomics Consortium epigenomes used for histone modification site enrichment testing. Metadata sourced from the Wang lab at Washington University in St. Louis (https://egg2.wustl.edu/roadmap/web_portal/meta.html).

| EID | Name | Type |
|---|---|---|
| E007 | H1 Derived Neuronal Progenitor Cultured Cells | ESCDerived |
| E009 | H9 Derived Neuronal Progenitor Cultured Cells | ESCDerived |
| E010 | H9 Derived Neuron Cultured Cells | ESCDerived |
| E054 | Ganglion Eminence derived primary cultured neurospheres | PrimaryCulture |
| E067 | Brain Angular Gyrus | PrimaryTissue |
| E068 | Brain Anterior Caudate | PrimaryTissue |
| E069 | Brain Cingulate Gyrus | PrimaryTissue |
| E071 | Brain Hippocampus Middle | PrimaryTissue |
| E072 | Brain Inferior Temporal Lobe | PrimaryTissue |
| E073 | Brain_Dorsolateral_Prefrontal_Cortex | PrimaryTissue |

| | | |
|---|---|---|
| E074 | Brain Substantia Nigra | PrimaryTissue |
| E125 | NH-A Astrocytes Primary Cells | PrimaryCulture |

**Table S3. Information on Allen Human Brain Atlas microarray donors.**

| Donor | Acb replicates | MFG-i replicates | Age window (yrs) | Sex | Ethnicity | PMI (hours) |
|---|---|---|---|---|---|---|
| 10021 | 7 | 13 | 36-40 | M | Black or African American | 10 |
| 12876 | 2 | 3 | 56-60 | M | White | 26 |
| 14380 | 2 | 4 | 31-35 | M | White | 17 |
| 15496 | 2 | 5 | 46-50 | F | Hispanic | 30 |
| 15697 | 2 | 5 | 51-55 | M | White | 18 |
| 9861 | 2 | 15 | 20-25 | M | Black or African American | 23 |

*All supplemental tables in Excel format can be found on figshare at doi: 10.6084/m9.figshare.24871662.

**Table S4.** Primary EWAS results (separate Excel file named 2304_EWAS_results.xlsx). For the final set of probes for results interpretation, we provide all summary statistics from the primary within brain region EWASs and the meta-analysis.

**Table S5.** Ethanol sensitivity analysis results (separate Excel file named Ethanol_DLPFC_NAc_Sensitivity_Results.xlsx). For the 105 significant CpGs from the primary brain region EWASs and the meta-analysis, we provide summary statistics for testing those CpGs against ethanol toxicology status within AUD cases.

**Table S6.** KEGG and GO pathway enrichment test results (separate Excel file named KEGG_GO_Results.xlsx).

**Table S7.** Results of location-based enrichment tests (separate Excel file named Enrichment_Fisher_Tests.xlsx).

**Table S8.** Results from comparisons to previously published EWAS (separate Excel file named Lookup_Comparison_Results.xlsx).

**Table S9. Results of testing for enrichment in top 1% of results from this study, Zillich et al. and Clark et al.**

| Prior analysis | DLPFC (BA 46/9) | | NAc | | Meta-analysis | |
|---|---|---|---|---|---|---|
| | N CpGs[1] | P-value[2] | N CpGs | P-value | N CpGs | P-value |
| Zillich - ACC | 636,087 | 0.5550 | 637,381 | 0.4557 | 634,959 | 0.3027 |
| Zillich - BA9 | 636,087 | 0.2601 | 637,381 | 0.4777 | 634,959 | 0.2795 |
| Zillich - CN | 671,801 | 0.6790 | 673,126 | 0.2842 | 670,631 | 0.3948 |
| Zillich - PUT | 671,637 | 0.1661 | 672,952 | 0.0398 | 670,468 | 0.2803 |
| Zillich - VS | 671,801 | 0.3898 | 673,126 | 0.0184 | 670,631 | 0.5163 |

| | | | | | | |
|---|---|---|---|---|---|---|
| Clark – BA10 hmCG | 653,348 | 0.1530 | 653,665 | 0.2199 | 652,424 | 0.3391 |
| Clark – BA10 mCG | 462,681 | 0.9966 | 462,903 | 0.5414 | 462,041 | 0.4746 |

[1]N CpGs represents the intersection of tested CpGs for each comparison of results, from which the top 1% of results were obtained.
[2]The permuted p-value, based on 10,000 permutations, for Cramer's V coefficient comparing the two sets of results.

**Table S10**. Results from LDSC analysis on varying genomic windows around significant CpGs (separate Excel file named LDSC_results.xlsx)

**Table S11.** Mean and standard deviation of meta-analysis $I^2$ statistics across significance groups.

| **Significance group** | **N probes** | **Mean (sd) $I^2$** |
|---|---|---|
| *DLPFC* | 26 | 89.6 (5.4) |
| *DLPFC & meta-analysis* | 5 | 67.1 (15.1) |
| *Meta-analysis* | 21 | 4.4 (11.1) |
| *NAc* | 48 | 88.1 (5.8) |
| *NAc & meta-analysis* | 5 | 57.4 (28.8) |

**Table S12.** Results from linear mixed-effects model with AUD, brain region, and brain region × AUD interaction as well as differentially methylated region (DMR) results for these same terms (separate Excel file named Cross_region_probe_dmr_results.xlsx).

**Table S13.** Results from comparing Allen Human Brain Atlas expression profiles of MFG-i and Acb brain regions using 1,000 bootstrap iterations of paired sample t-tests, for genes annotated to significant DMRs from the linear mixed-effects modeling analysis (separate Excel file named Cross_region_AHBA_results.xlsx).

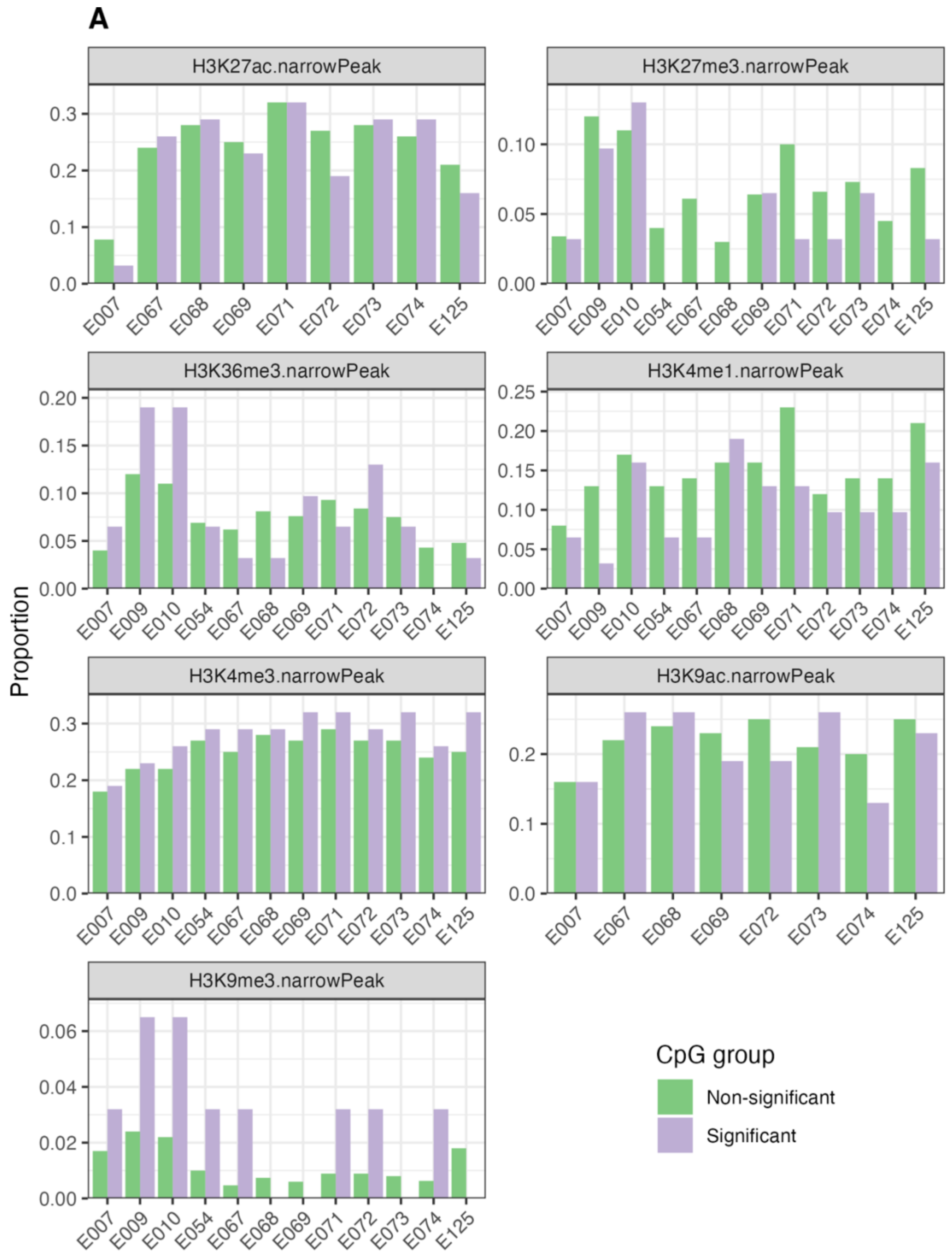

A
H3K27ac.narrowPeak
H3K27me3.narrowPeak
H3K36me3.narrowPeak
H3K4me1.narrowPeak
H3K4me3.narrowPeak
H3K9ac.narrowPeak
H3K9me3.narrowPeak
Proportion
E007 E009 E010 E054 E067 E068 E069 E071 E072 E073 E074 E125
CpG group
Non-significant
Significant

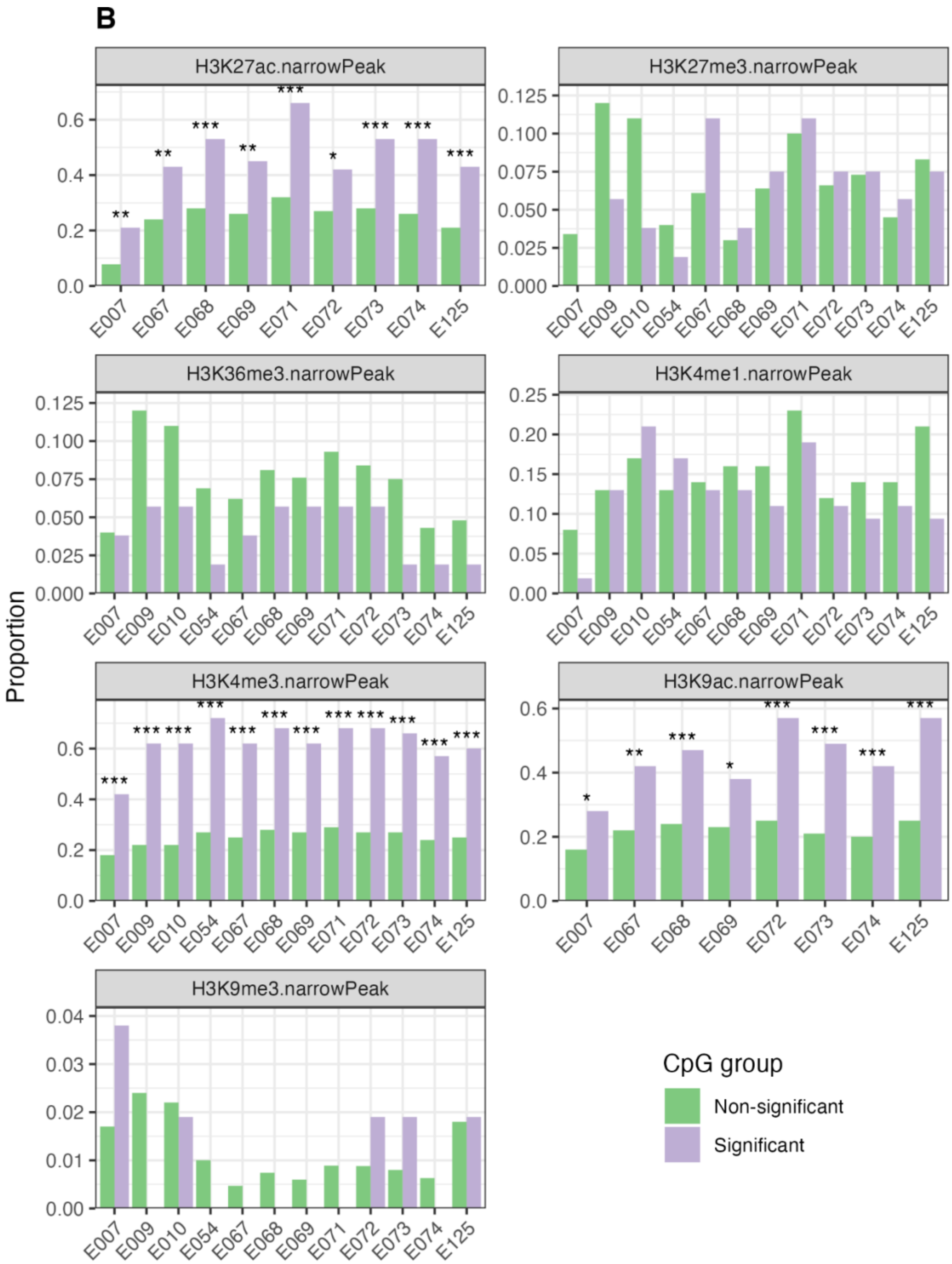
B
H3K27ac.narrowPeak
H3K27me3.narrowPeak
H3K36me3.narrowPeak
H3K4me1.narrowPeak
H3K4me3.narrowPeak
H3K9ac.narrowPeak
H3K9me3.narrowPeak
Proportion
E007
E009
E010
E054
E067
E068
E069
E071
E072
E073
E074
E125
CpG group
Non-significant
Significant

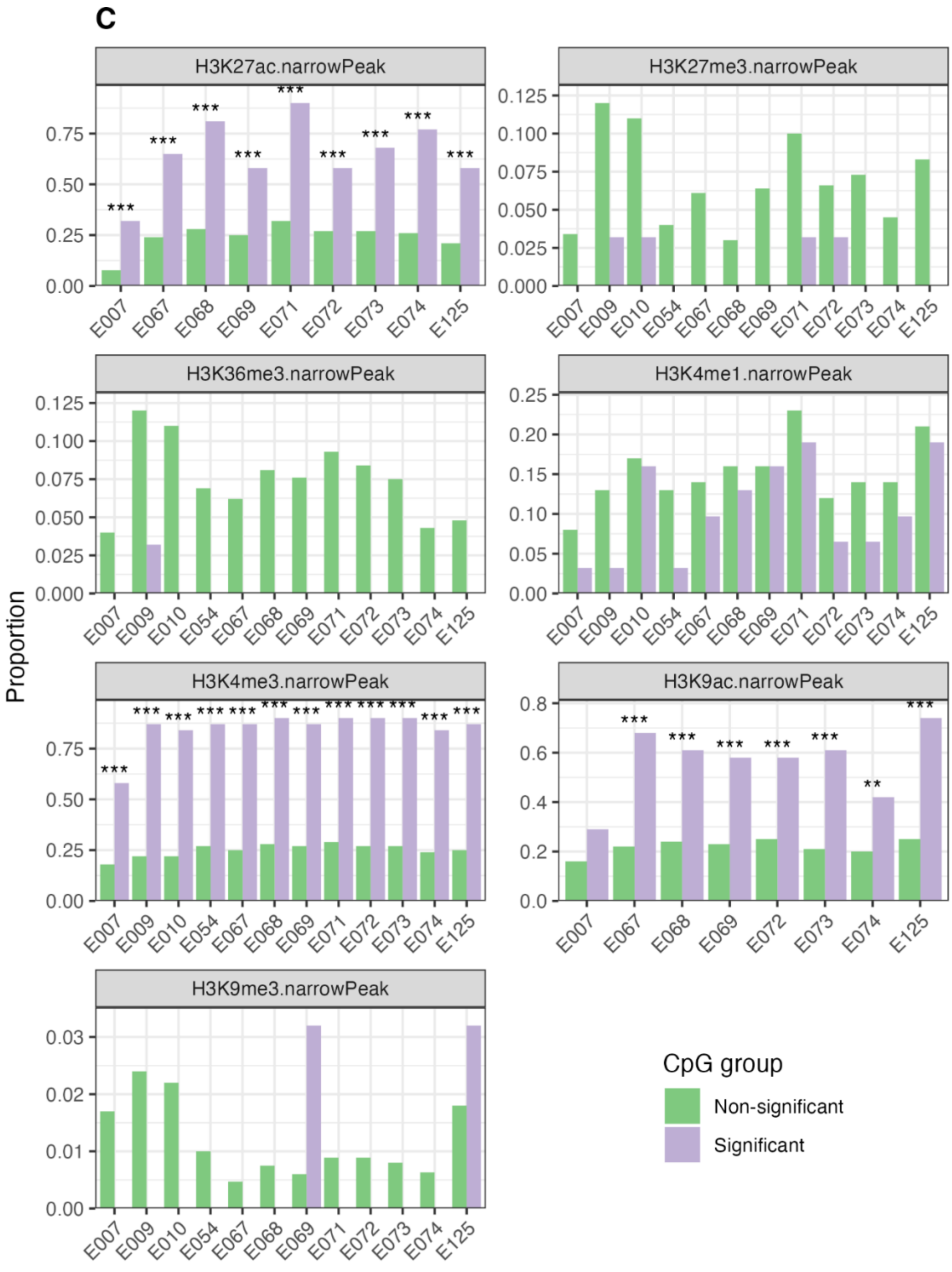


**Figure S1. Enrichment of histone modification sites.** For CpGs from the DLPFC EWAS (A), the NAc EWAS (B), and the meta-analysis (C), the proportions of non-significant vs. significant CpGs were compared based on ChIP-seq histone modification marks from the Roadmap Epigenomics

Consortium. Stars represent significance of Fisher's exact test for count data, based on a two-sided test with FDR-corrected p-values; p < 0.001 is ***, p < 0.01 is **, and p < 0.05 is *.

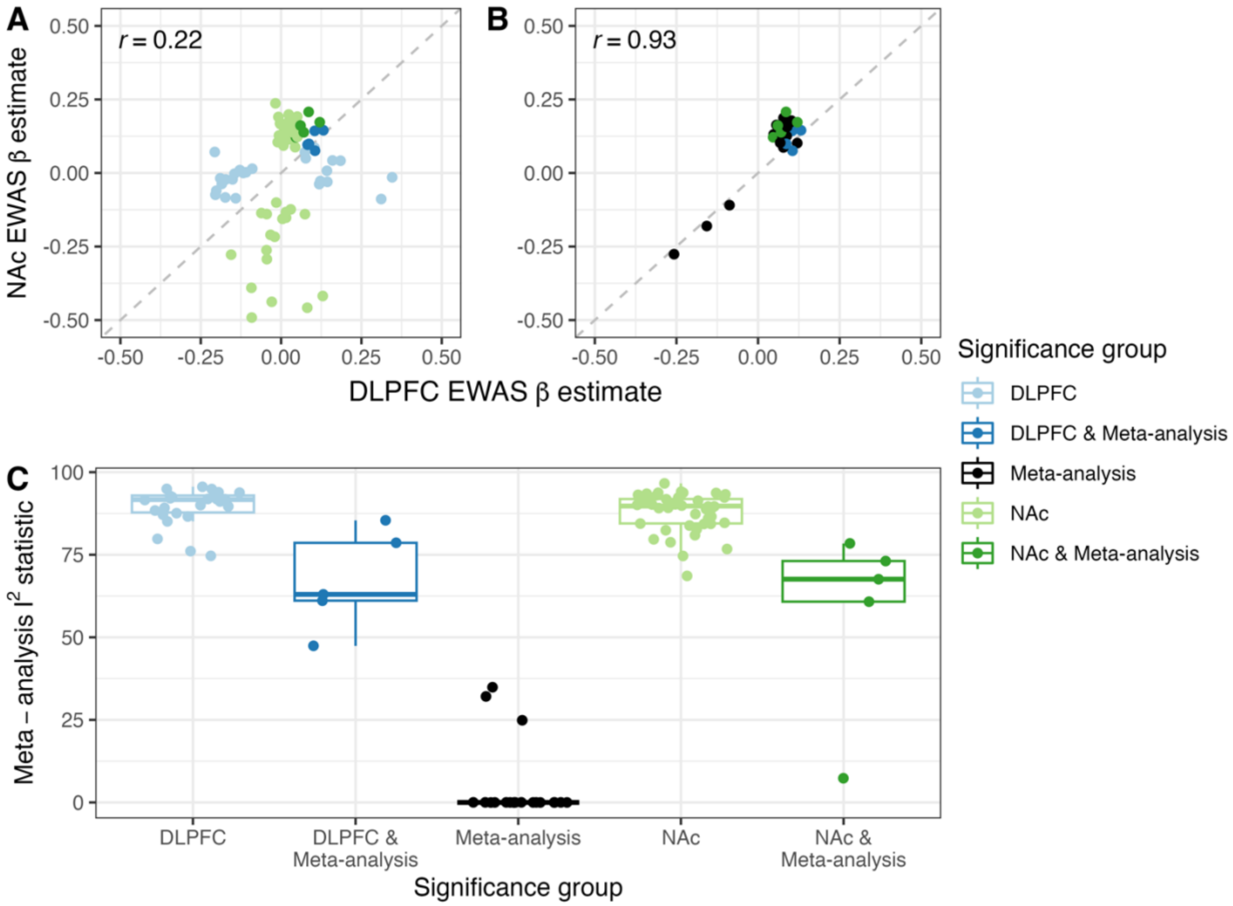


**Figure S2. Comparison of effects and $I^2$ heterogeneity statistics across significance groups.** Top row of plots contains a comparison of M-value EWAS ß estimates from DLPFC and NAc specific analyses, colored by the analysis in which the probe reached the FDR significance threshold. Probes that were significant in analyses of DLPFC or NAc regions are in (A), and probes that were significant in the meta-analysis are in (B). Note there are 13 probes represented in both plots, as they were significant in the meta-analysis and one of the brain region analyses. The bottom plot (C) compares meta-analysis $I^2$ statistics across the same set of significance groupings.

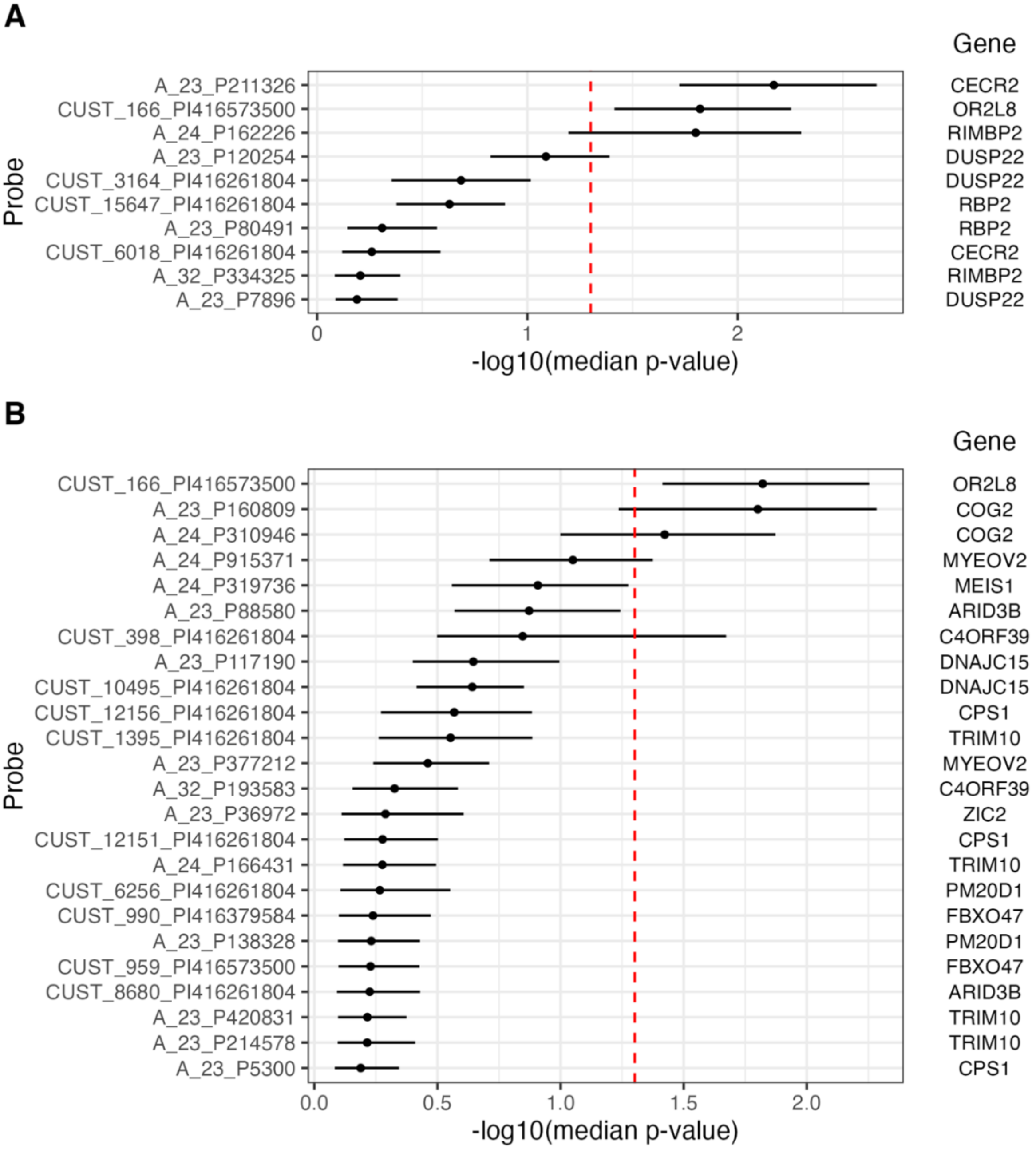


**Figure S3. Comparison of MFG-i and Acb expression profiles for genes annotated to nominally significant DMRs from the linear mixed effects modeling.** Using the Allen Human Brain Atlas data from MFG-i (prefrontal cortex) and Acb (nucleus accumbens), we conducted a paired-sample t-test to compare expression profiles in these two brain regions for genes that were annotated to differentially methylated regions (DMRs) reaching FDR significance in our linear mixed effects modeling for the brain region term (A) and the interaction term (B).
